# The association between Internet Usage and Overweight/Obesity modified by Gender: Evidence from a nationally representative survey in Nepal

**DOI:** 10.1101/2020.07.18.20156661

**Authors:** Juwel Rana, Md. Momin Islam, John Oldroyd, Nandeeta Samad, Rakibul M. Islam

## Abstract

**Objective:** Using a nationally representative data, we examined the associations between internet use and overweight/obesity in people aged 15-49 years in Nepal, and the extent to which these associations vary by gender.

**Materials and methods:** The study analyzed the nationally representative Nepal Demographic and Health Survey (NDHS) 2016 data, collected between June 2016 and January 2017. The outcome was overweight/obesity. Exposures were internet use (IU) in the last twelve months and internet use frequency (FIU) in the last month. Multivariable ordinal logistic regression models were fitted to estimate the total effects of IU and FIU on overweight/obesity adjusted for minimal sufficient adjustment set of potential confounders. P-difference was extracted using a Wald test for the models with interaction terms.

**Results:** Of the 10,380 participants, 33.9% used internet in the last 12 months, and 13.1 % used less than/at least once in a week, and 17.5% used internet almost every day. The prevalence of overweight/obesity by IU was 38% (95% CI: 35.9%, 40.1%) for male and 44.1% (95% CI: 41.6%, 46.6) for female. The risk of overweight and obesity was significantly 1.55 times higher (aOR: 1.55; 95% CI: 1.40, 1.73; p < 0.001) among those participants who used the internet compared to the individual who did not use the internet in the last 12 months or earlier of the interview. Similar associations were observed when using the augmented measure of exposure-FIU. We observed modification effect of gender in the associations of IU (p-difference<0.001) and FIU (p-difference<0.002) with overweight and obesity in Nepal.

**Conclusions:** Our findings suggest that it is imperative for future overweight/obesity interventions in LMICs, including Nepal, to discourage unnecessary internet use, particularly among males.

## Introduction

Overweight/obesity- a major risk factor for non-communicable disease (NCD), premature deaths, and disability- is increasing exponentially worldwide, especially among people in low and middle-income countries (LMICs) [1,2]. Physical inactivity and sedentary lifestyles significantly contribute to this trend [3]. The internet has become an integral source of daily communication and information in contemporary society with its easy accessibility, cost-effectiveness, and time-saving attributes [4]. This increasing dependency on the internet has an apparent impact on sedentary lifestyle and physical inactivity [5,6], which is associated with overweight/obesity and related diseases and complications [7,8].

Studies have shown that excessive internet use and internet-based activities, especially during weekends and leisure hours, affect individuals’ regular food consumption patterns, standard sleep hours, and sleeping schedules that are associated with weight gain [9–11]. One study showed that night-time internet use, especially for video games and complementary day-time sleeping, is significantly associated with increased body mass index (BMI) [12]. Numerous studies have also found that prolonged internet usage (3 hours or more a day) and internet-based recreational activities, including watching online movies, reading newspapers, playing games, and watching television, are positively associated with BMI among adolescents and adults [13–16]. In addition, there is evidence that greater use of electronic devices for online gaming and social networking leads to increased consumption of sugar-sweetened beverages [17] and, therefore, overweight and obesity [18].

Overweight/obesity prevalence is higher in women than in men, including in Nepal [19,20], due to several biological factors, such as fat distribution and hormonal influences (19). Socio-cultural factors also significantly contribute to this hypothesis due to leisure hours as well as non-demanding activities [21,22]. In relation to internet use, there is evidence of a “digital divide” between male and female users of the internet [23] This is characterized by the internet predominantly being accessed by males rather than females due to societal roles, occupation, economic factors that lead to differential health consequences [24]. As a further matter, due to technophobia and absence of internet literacy, older people, particularly older women, are reluctant to use the internet. In contrast, the younger generation, irrespective of sex, use the internet for a variety of purposes [23].

The prevalence of overweight/obesity and internet use are increasing in Nepal. Studies have found that the prevalence of overweight/obesity has increased from 21% in 2015 to 31% in 2018 in Nepal [19,20] while the prevalence of internet use has increased from 9% in 2011 to 63% in 2019 [25]. We hypothesized that an increasing trend of overweight/obesity in Nepal could be explained by an increasing use of internet that has not been explored yet. To address this knowledge gap, using a nationally representative data, we examined the associations between internet use and overweight/obesity in people aged 15-49 years in Nepal, and the extent to which these associations vary by gender.

## Materials and Methods

### Data source and study setting

The study analyzed the nationally representative Nepal Demographic and Health Survey (NDHS) 2016 data, collected between June 2016 and January 2017.

### Study design and study populations

Data on this study were obtained from the NDHS 2016 implemented by New ERA under the supervision of the Ministry of Health, Nepal. The households of the NDHS 2016 were selected in two ways based on the urban/rural locations. Firstly, the two-stage stratified sampling process was used in rural areas where wards were selected in the first stage as a primary sampling unit (PSUs), and households were selected in the second stage. Secondly, three-stage stratified sampling was used in urban areas to select households where wards were selected in the first stage (PSUs), enumeration areas (EA) were selected from each PSU in the second stage and households were selected from EAs in the third stage. A total of 383 wards, 184 urban and 199 rural, were selected all together. Finally, a total of 11,490 households (rural-5,970 and urban-5,520) were selected for the NDHS 2016 [26]. Data were collected by the trained interviewers visiting the households. The overall response rate was approximately 97%. BMI was measured among 14,763 individuals with 6,120 men and 8,647 women aged 15-49 years. In our study, a total unweighted sample was 10,380 comprising men (3,995) and women (6,385), after excluding participants aged >49 years and discarding the missing and extreme values. The total weighted analytic sample was 10,384 participants (men 4,054 and women 6,330) aged 15-49 years. Details of the NDHS 2016, including survey design, sample size determination, and questionnaires, have been described elsewhere [26].

### Measures of outcomes: Body mass index

The outcome of interest in this study BMI was categorized using both South Asia (SA) specific and World Health Organization (WHO) cut-off. According to SA cutoff values, underweight was defined as BMI <18.50 kg/m^2^, normal 18.50 kg/m^2^oto <23.00 kg/m^2^, overweight 23.00 kg/m^2^ to 27.50kg/m^2^ and obesity was defined as BMI ≥27.50 kg/m^2^. According to WHO, the corresponding categories were underweight (<18.50 kg/m^2)^, normal (18.50 kg/m^2^ to<25.00 kg/m^2^), overweight (25.00 kg/m^2^ to 29.99 kg/m^2^) and obesity (≥ 30.00 kg/m^2^).

### Measures of exposures: Internet usage

Internet use (IU) in the last twelve months and internet use frequency (FIU) in the last month were two main exposures of interest. IU was categorized as (1) no and (2) yes, while FIU was categorized as (1) non-user, (2) less than a week/at least once in week, and (3) almost every day.

### Covariates and potential confounders

A number of covariates have been selected in this study based on prior literature and researchers’ subject knowledge [27–30]. These are age group (15-24, 25-34, 35-44 and 45-49), gender (male and female), urbanicity (urban and rural), ecological zone (Mountain, Hill, and Terai), education (no education, primary, secondary and higher), household wealth quintiles (poorest, poorer, middle, richer and richest), number of under-five children (no, one, two, and 3+), occupations (unemployed, non-manual job, manual job, and agriculture), marital status (unmarried and ever married including divorced, widow and separated), watching television (TV) (not at all, less than once a week and at least once a week), caffeine (no and yes) and current tobacco use (yes, no).

### Statistical Analysis

We first present the characteristics of the study population by BMI categories and the exposure variables (IU and FIU). The univariate associations of BMI categories and the internet usages with different sample characteristics were examined by chi-square tests. Multivariable ordinal logistic regression models were fitted to estimate the total effects of IU and FIU on overweight/obesity adjusted for minimal sufficient adjustment set of potential confounders based on the DAG model (Fig 2 and Table 1), and effect modification of gender in these associations. To avoid multicollinearity and over adjustment, we did not adjust for watching TV and level of education. Results were presented as adjusted odds ratio (aOR) and 95% confidence interval (CI). All analyses accounted for complex survey design to adjust clustering variation, the probability of selection, and non-response in the NDHS [31]. All statistical tests were two-sided, and a p-value <0.05 was the level of significance. *P*-difference was extracted using a Wald test for the models with interaction terms. Analyses were conducted using software R version 4.2 [32].

**Fig 1.**
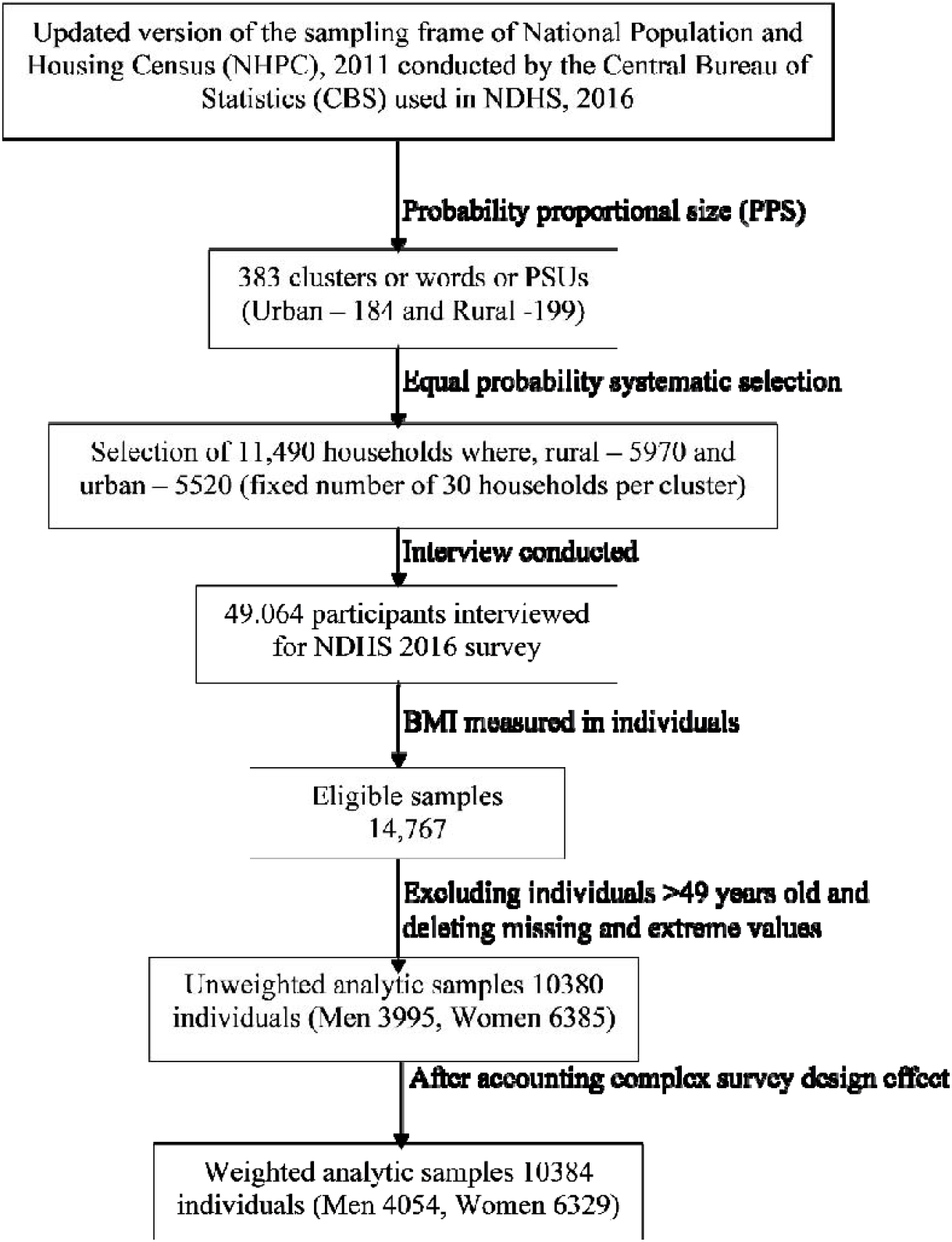
Flowchart of the analytic sample selection process.

**Table 1.** **The weighted prevalence of underweight, normal weight, and overweight/obesity (South Asian Cut-off) by sample characteristics in Nepal 2016**

| Characteristics | Sample Distribution (%) | Prevalence of (%) underweight, normal weight, and overweight/obesity |  |  |  |
| --- | --- | --- | --- | --- | --- |
|  |  | Under weight | Normal | Over weight/ Obesity | P-value |
| Overall | 10380 (100) | 16.5 (1722) | 48.5 (5243) | 35.0 (3415) | - |
| Age group |  |  |  |  |  |
| 15-24 | 4048 (38.2) | 25.1 | 57.1 | 17.8 | <0.001 |
| 25-34 | 2950 (29.0) | 11.7 | 46.5 | 41.8 |  |
| 35-44 | 2417 (23.7) | 10.7 | 39.3 | 50.0 |  |
| 45-49 | 965 (9.1) | 11.0 | 43.4 | 45.6 |  |
| Gender |  |  |  |  |  |
| Male | 3995 (39.0) | 16.8 | 50.7 | 32.5 | <0.001 |
| Female | 6385 (61.0) | 16.3 | 47.1 | 36.6 |  |
| Marital status |  |  |  |  |  |
| Unmarried | 2609 (25.4) | 27.8 | 56.9 | 15.3 | <0.001 |
| Ever Married | 7771 (74.6) | 12.7 | 45.7 | 41.6 |  |
| Number of under five years old child |  |  |  |  |  |
| No under 5 | 5892 (57.0) | 15.5 | 47.7 | 36.8 | <0.001 |
| One | 2910 (27.9) | 16.0 | 48.5 | 35.5 |  |
| Two | 1241 (11.9) | 20.6 | 51.8 | 27.6 |  |
| 3+ | 337 (3.2) | 23.4 | 52.4 | 24.2 |  |
| Educational qualification |  |  |  |  |  |
| No education | 2524 (24.0) | 17.8 | 50.7 | 31.5 | <0.001 |
| Primary | 1783 (17.7) | 17.3 | 46.1 | 36.6 |  |
| Secondary | 4325 (40.9) | 17.8 | 49.0 | 33.2 |  |
| Higher | 1748 (17.4) | 10.8 | 47.0 | 42.2 |  |
| Occupational status |  |  |  |  |  |
| Unemployed | 2579 (25.3) | 21.4 | 46.1 | 32.5 | <0.001 |
| Non-manual job | 2080 (21.7) | 8.3 | 39.9 | 51.8 |  |
| Agriculture | 4436 (40.0) | 17.4 | 54.2 | 28.4 |  |
| Manual job | 1285 (13.0) | 17.8 | 50.2 | 32.0 |  |
| Wealth index |  |  |  |  |  |
| Poorest | 2109 (16.6) | 18.6 | 59.5 | 22.0 | <0.001 |
| Poorer | 2088 (18.8) | 20.5 | 52.3 | 27.2 |  |
| Middle | 2106 (20.1) | 19.7 | 51.4 | 28.9 |  |
| Richer | 2204 (23.1) | 15.9 | 47.6 | 36.5 |  |
| Richest | 1873 (21.4) | 9.0 | 35.2 | 55.8 |  |
| Current Tobacco use (any type) |  |  |  |  |  |
| No | 7631 (73.9) | 17.0 | 47.5 | 35.5 | 0.014 |
| Yes | 2749 (26.1) | 15.1 | 51.6 | 33.3 |  |
| Coffee, tea, cola, or other drink (Caffeine) |  |  |  |  |  |
| No | 9809 (94.2) | 16.8 | 48.9 | 34.3 | <0.001 |
| Yes | 571 (5.8) | 10.9 | 43.0 | 46.1 |  |
| Frequency of watching television |  |  |  |  |  |
| Not at all | 2901 (25.7) | 21.5 | 54.0 | 24.5 | <0.001 |
| Less than once a week | 2644 (24.2) | 17.3 | 54.2 | 28.5 |  |
| At least once a week | 4835 (50.1) | 13.6 | 43.0 | 43.4 |  |
| Urbanicity |  |  |  |  |  |
| Urban | 6736 (63.0) | 15.5 | 46.0 | 38.5 | <0.001 |
| Rural | 3644 (37.0) | 18.3 | 52.8 | 28.9 |  |
| Ecological zone |  |  |  |  |  |
| Mountain | 751 (6.1) | 12.0 | 56.1 | 31.9 | <0.001 |
| Hill | 4668 (43.6) | 11.6 | 48.6 | 39.8 |  |
| Terai | 4961 (50.3) | 21.3 | 47.6 | 31.1 |  |
| Internet use (IU) at the last 12 months |  |  |  |  |  |
| No | 7134 (66.1) | 17.9 | 50.0 | 32.1 | <0.001 |
| Yes | 3246 (33.9) | 13.9 | 45.6 | 40.5 |  |
| Frequency of internet use (FIU) in the last 12 months |  |  |  |  |  |
| Non-user | 7472 (69.4) | 17.8 | 49.9 | 32.3 | <0.001 |
| Less than/at least once in a week | 1324 (13.1) | 16.4 | 47.0 | 36.6 |  |
| Almost every day | 1584 (17.5) | 11.4 | 44.5 | 44.1 |  |

**Fig 2.**
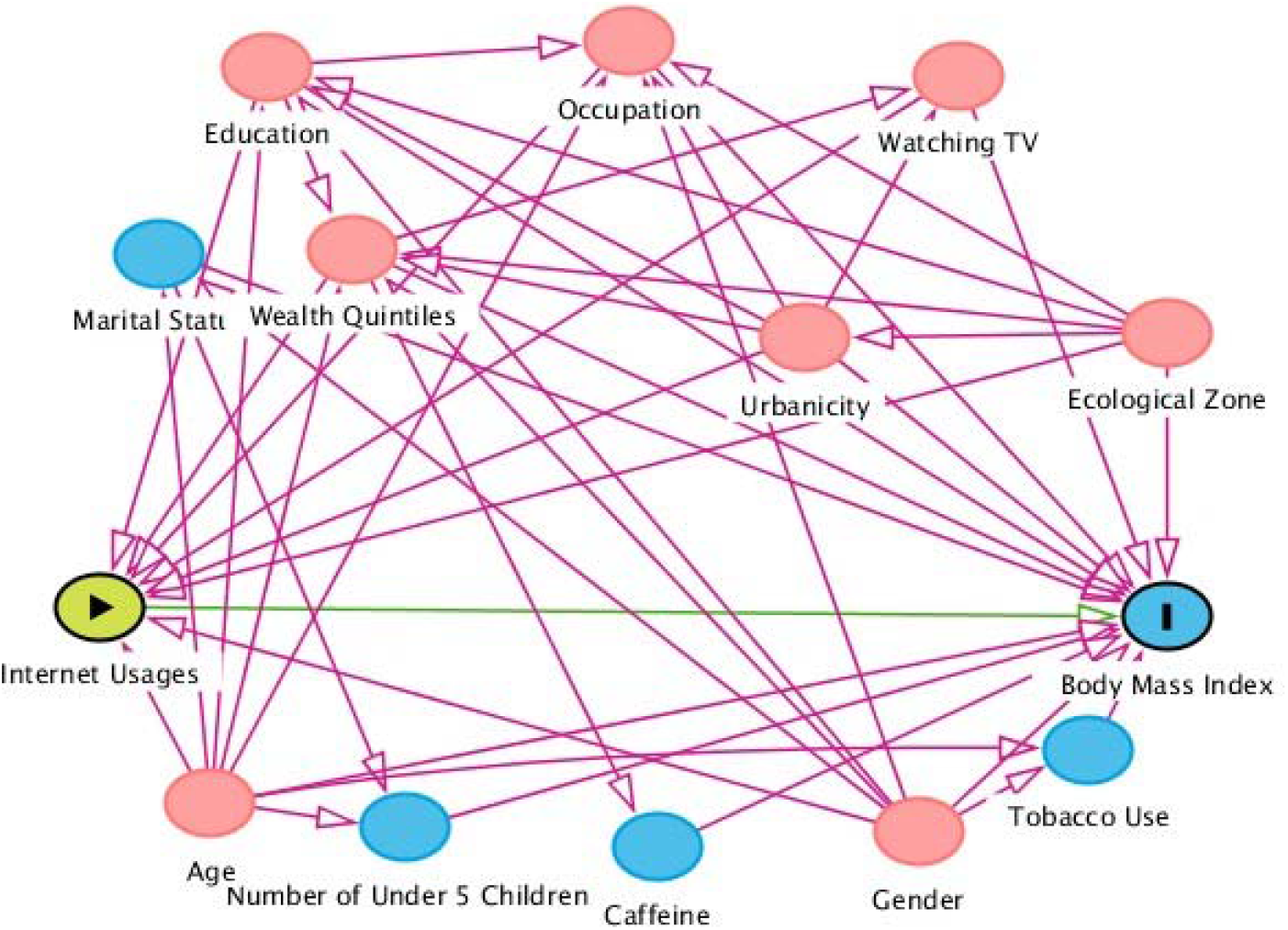
A directed acyclic graph (DAG) for adjusting confounders in the associations between internet usages and overweight/obesity. Overweight/obesity was the outcome variable, and internet usages were the exposure variable. Minimal sufficient adjustment sets for estimating the total effect of internet usages on BMI contains age, ecological zone, education, gender, occupation, urbanicity, watching TV, and wealth quintiles. This figure was constructed through DAG (Online access: http://dagitty.net/mVo4Gb9).

### Data and Code Availability

All data files are available from the DHS program database: https://dhsprogram.com/data/dataset/Nepal_Standard-DHS_2016.cfm. Codes are available upon request.

## Results

### Sample characteristics

A total of 10380 (unweighted sample) participants were included in the analysis, and their characteristics (weighted percentage) are presented in Table 1. The mean (±SE) age of the participants was 29.4 (*±*0.11). Of the study participants, 61% were female, 74.6% were either married or ever married that included divorced, separated, or widow. About two-thirds (63%) of the participants lived in urban areas, while about half of them (50.1%) watched television at least once in a week. About one-quarter (24.0%) of the participants had no education, 40% of the participants were involved in agricultural work, and 35.4% of the participants were from the poorest and poorer households.

### Weighted prevalence of overweight/obesity and internet usages by sample characteristics

The univariate associations of overweight/obesity and internet usages are presented in Table 1, sTable 1, and Fig 3. The overall prevalence of overweight/obesity was 35.0% (Table 1).

**Fig. 3:**
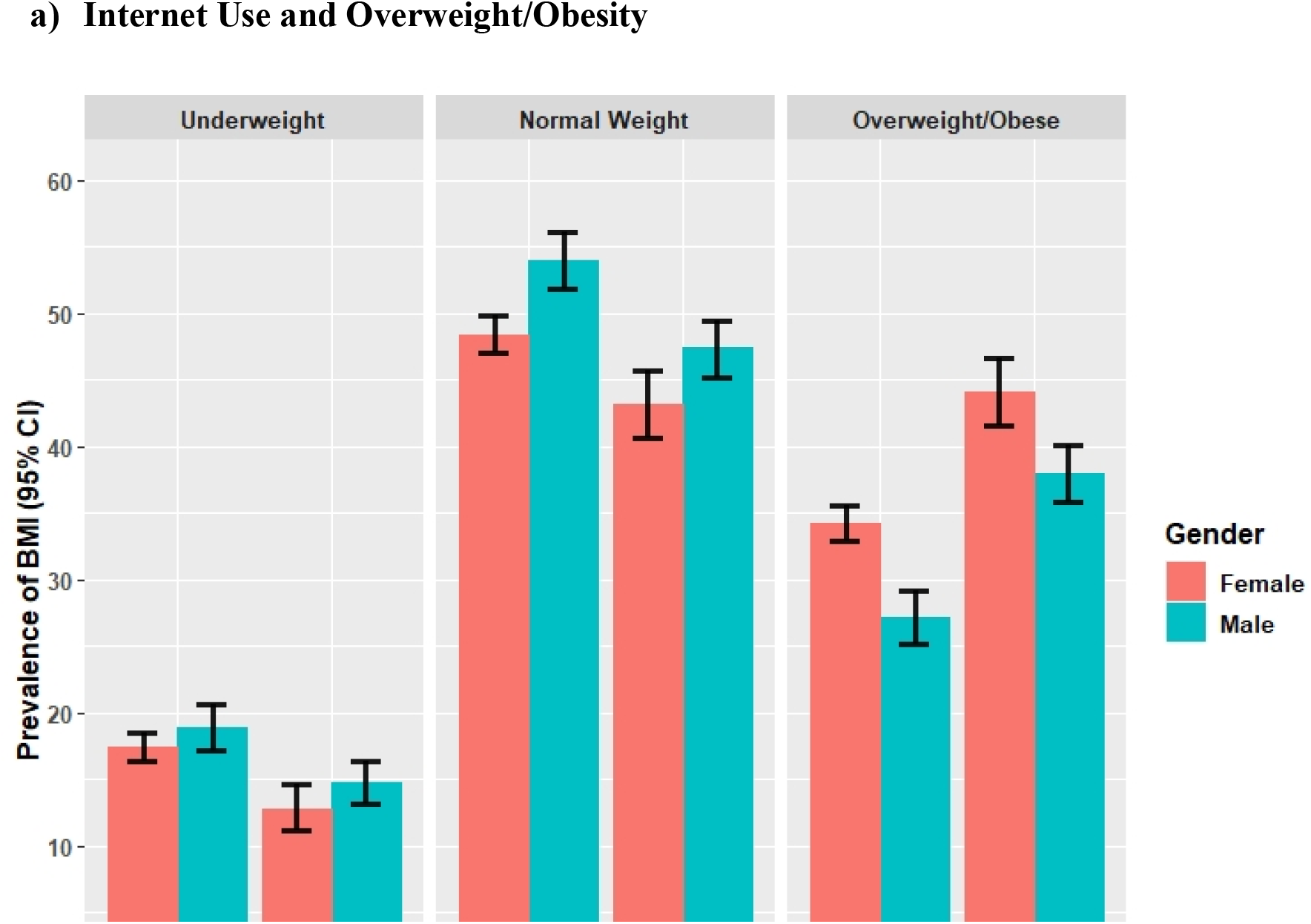
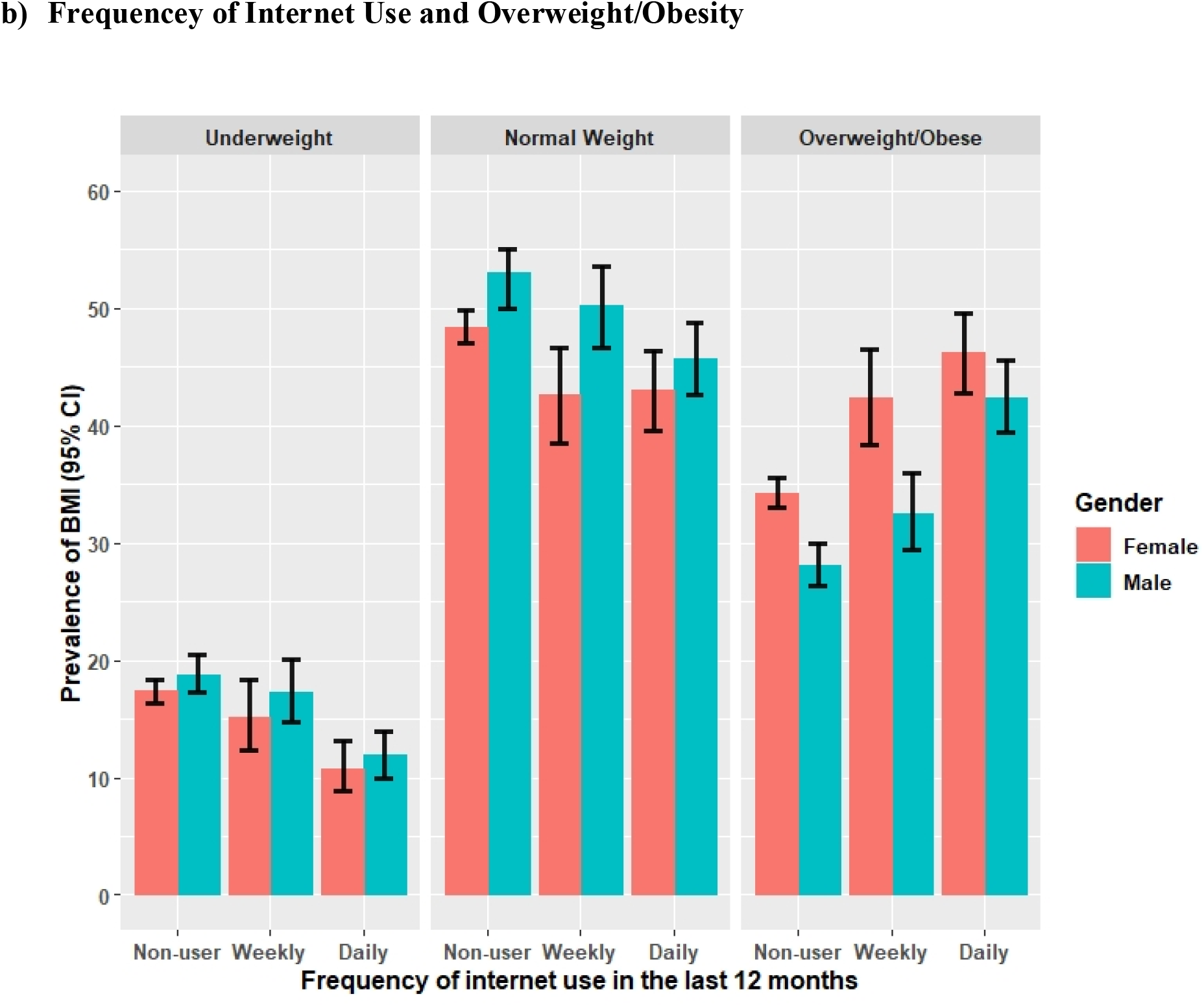
Gender Stratified Weighted Prevalence of Underweight, Normal Weight and Overweight/Obesity by Internet Usages.

The gender stratified weighted prevalence of BMI categories by internet usages were shown in Fig 3. The prevalence of overweight/obesity by IU was 38.0% (95% CI: 35.9%, 40.1%) for male and 44.1% (95% CI: 41.6%, 46.6) for female, while the corresponding prevalence of overweight/obesity were 32.5% (95% CI: 29.4, 36.0%) for males and 42.4% (95% CI: 38.4%, 46.5%) who used internet at least once in week, and 42.4% (95% CI: 39.4%, 45.6%) for males and 46.2% (95% CI: 42.8%, 49.6%) who used internet almost every day.

Table 1 also shows that the prevalence of overweight/obesity was highest among those who were ever married (41.6%), highly educated (42.2%), non-manual job holders (51.8%), richest (55.8%), weekly television viewers (43.4%), urban residents (38.5%) and residents living in Hill (39.8%).

Of the 10,380 participants, 33.9% used internet in the last 12 months, and 13.1 % used less than/at least once in a week, and 17.5% used internet almost every day (sTable 1). The prevalence of IU among male (49.4%) was higher compared to the female (23.9%). People with higher education, wealth quintiles, and non-manual jobs had higher percentages of internet usages.

### Multivariable associations between internet usages and overweight/obesity

The ordered logistic regression models were developed to estimate the total effect of internet usages on overweight/obesity after adjusting for age, gender, marital status, occupation, household wealth index, urbanity, and ecological zone and the effect modification of gender in the associations between internet usages and overweight/obesity (Fig 4). Results show that the risk of overweight and obesity was significantly 1.55 times higher (aOR: 1.55; 95% CI: 1.40, 1.73; p < 0.001) among those participants who used internet compared to the individual who did not use internet in the last 12 months or earlier of the interview. When using the augmented measure of exposure-FIU, the risk of overweight and obesity was significantly 1.51 times higher (aOR: 1.55; 95% CI: 1.31, 1.74; p < 0.001) among those participants who used the internet less than/ at least once in a week compared to the participants who did not use the internet. Similarly, the participants who used internet almost every day (aOR: 1.56; 95% CI: 1.35, 1.79; p < 0.001) had 1.56 times higher risk of being overweight and obese compared to participants who did not use internet (Fig 4).

**Fig 4:**
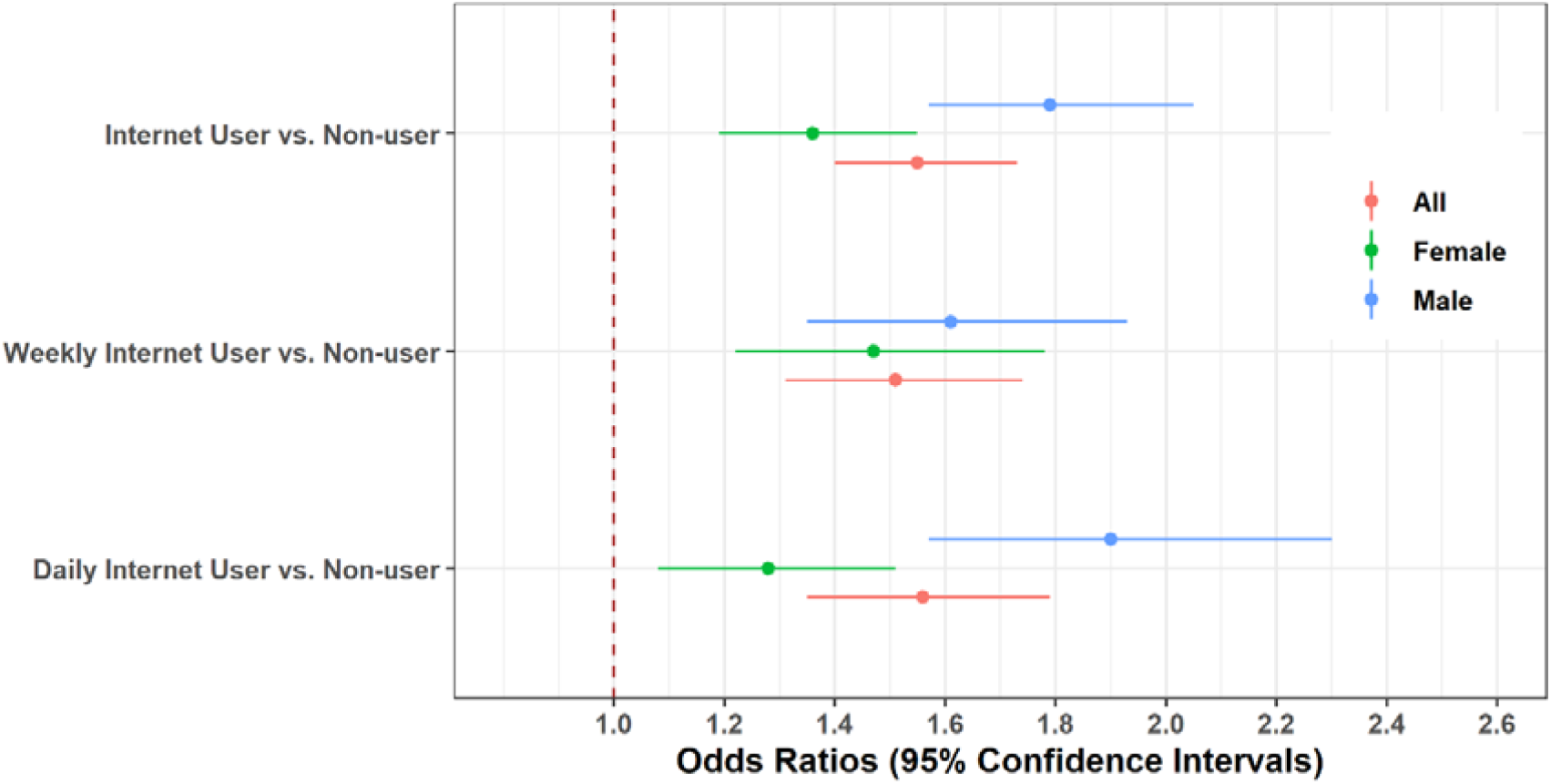
Associations between internet usages and overweight/obesity (South Asian Cut-off) modified by gender in Nepali adult populations 2016. ^a^Ordered logistic models without interaction term were adjusted for age, gender, marital status, occupation, household wealth index, urbanicity, and ecological zone.

When exploring the effect modification by gender, we observe that the risk of overweight and obesity were significantly 1.79 (aOR: 1.79; 95% CI: 1.57, 2.05) times higher among males who used internet in the last 12 months or earlier compared to males who did not use internet. A similar association was observed among females who used internet (aOR: 1.36; 95% CI: 1.19, 1.55). However, the differences of effect estimates were significantly different (*p*-difference<0.001) between males and females. Likewise, we observed modification effect of gender in the association between FIU and overweight and obesity in Nepal (Fig 4). In other words, the risk of being overweight and obese was higher among males compared to females (p-difference=0.002) who used internet frequently.

### Sensitivity analyses

As a part of the sensitivity analyses, an additional analysis was conducted using global cut-off of BMI categories, and we found similar results (data not shown).

## Discussion

This is the first study to investigate an association between internet use and overweight/obesity and the moderating effects of this association, a novel finding, in a LMIC. Using data from a large, nationally representative survey in Nepal, we have found that in adjusted analysis, the risk of overweight and obesity was significantly higher among those people who used the internet compared to people who did not use internet in the last 12 months. This was also true among those participants who used the internet less than/ at least once in a week or who used internet almost every day, compared to the participants who did not use the internet. We also found effect modification by gender - the risk of overweight and obesity was higher among males compared to females who used the internet frequently. These data have implications for public health measures to control overweight and obesity in LMIC.

After adjusting for age, gender, marital status, occupation, household wealth index, urbanicity, and ecological zone, our study showed that internet use, either less than/at least once in a week or almost every day, is independently and positively associated with increased odds of being overweight and obese. This is consistent with a recent systematic review and meta-analysis of nine cross-sectional studies from high and LMICs [8]. Furthermore, excessive internet use during weekends and leisure hours and/or frequency of internet use has a positive association with overweight and obesity [8,33], which is in agreement with our study. The widespread uptake of internet use across the world may be contributing to the rising rate of overweight and obesity globally [34,35] an observation which is supported by our study.

It is a matter of great concern that the increased dependency on the internet has initiated an apparent drift from physical mobility to sedentary lifestyle [33,36]. Consistent with this, one of the greatest changes in the last decade in Nepalese lifestyle is that internet use has increased substantially in more than half of the total population [25]. Possible mechanisms through which internet use may cause obesity include a disrupted sleeping schedule leading to weight gain as well as effects on an individuals’ regular food consumption [10,11,37]. Increasing consumption of snacks and sugar-sweetened beverage consumption during online gaming or watching movies online [18] may also result in excess fat accumulation [17]. In addition, increasing computer usage contributes to obesity in households in other LMICs, including China, Ghana, and South Africa [38–40].

We explored the modification effect of gender on internet use and obesity. We found the risk of overweight/obesity was significantly higher among males who used internet in the last 12 months or earlier compared to those who did not. A similar but weaker association was observed among females. This statistically constructed evidence on the effect of gender modification on internet use frequency and its association with overweight/obesity suggests that males who are exposed to internet users have higher obesity and overweight rate than females. Notwithstanding the modification effects we have found of gender on internet use and obesity, the findings underline that internet use is relevant to obesity in both males and females. There are multiple lifestyle influences on obesity, of which internet use is only one. Our data provide clear preliminary signals that internet use does need to be considered in obesity interventions in both genders.

Whereas previous studies have examined the association between television watching and obesity [41], examining internet use and obesity is a new finding, particularly in LMICs. Television viewing is a proxy for physical inactivity and has shown an association with numerous risk factors, including poor diet, lower socioeconomic status, obesity, smoking, or depressive symptoms [41]. Whereas television viewing is more likely to occur for relaxation, internet use is greater as it can be either for relaxation (e.g., playing games, social media participation, online movies, reading newspapers and other internet-based recreational activities,) or work (use of libraries, accessing work-related websites, email, banking, appointments, meeting and other online work activities). Both uses are likely to result in an increase in physical inactivity. Internet use, however, is a more contemporary proxy for physical inactivity. In future, direct measures of ‘weekly screen time’ downloaded from internet-enabled devices rather than recalled estimates will be able to be used to examine the associations between internet use and obesity with greater precision.

The major strengths of our study include a large, nationally representative sample from survey in Nepal and using robust statistical methods to minimized bias (e.g., directed acyclic graph (DAG) to adjust for confounders). Importantly, we examined the association between internet use and overweight/obesity using both South Asian specific and WHO cut-offs. Our findings were consistent across both measures. Limitations include that inferences about causality cannot be made due to cross-sectional data. In fact, the relationship between internet use and obesity may be bi-directional. Future prospective studies will be able to determine the strength and direction of the causal association between internet use and obesity. We adjusted for all confounding variables available to us (e.g., socioeconomic status, television viewing, and current tobacco use); however, other relevant variables were not collected as part of the survey (e.g., dietary intake and physical activity). Although this does not materially alter our main findings, it weakens the precision of the estimates of association we have observed. We measured internet use as exposure in two ways: internet use in the last twelve months and an augmented measure of internet use frequency in the last month. It is possible that both measures are subject to recall bias. However, our results are consistent across both measures. Further, we observed a tendency for greater overweight/obesity with more frequent internet use using either weighted prevalence obesity estimates or logistic regression. These results underline our key finding that the risk of overweight and obesity was significantly higher among those individuals who used internet compared with individuals who did not use internet. Whether there is a dose-response effect of internet use on overweight/obesity-more internet use leads to greater overweight/obesity, as suggested by our results-need further detailed investigation.

## Conclusion

In conclusion, internet use is independently and positively associated with overweight/obesity in Nepal. A comprehensive assessment of how gender interacts with internet usage and obesity suggests that the risk of overweight and obesity was higher among males compared with females in Nepal who used the internet frequently. Our findings suggest that it is imperative for future overweight/obesity interventions in LMICs, including Nepal, to discourage unnecessary internet use, particularly among males.

## Data Availability

All data files are available from the DHS program database: https://dhsprogram.com/data/dataset/Nepal_Standard-DHS_2016.cfm.

## Acknowledgments

The authors are grateful to MEASURE DHS for granting access to the Nepal Demographic and Health Survey 2016 Data.

## Authors’ contribution

JR and RI developed the study concepts. JR and RI analyzed the data. JR, MMI, JO, and NS drafted the manuscript. All authors read, critically reviewed, and approved the final version of the paper.

## Ethical approval and consent to participate

The Nepal Health Research Council and the ICF Institutional Review Board (IRB) and the Ministry of Health Ramshah Path, Kathmandu, Nepal, approved the NDHS 2016 survey protocol. The 2016 NDHS required written consent from the household head to carry out the interviews and anemia testing. Informed consent was taken from each participant before the enrollment. For the current study, we obtained approval from ICF in June 2018 to use the de-identified data of the DHS online archive.

## Supporting Information

### Supplementary Table

sTable 1 The weighted prevalence of internet use and frequency of internet use in the last 12 months by sample characteristics in Nepal 2016.

## References

1. Wang Y, Beydoun MA, Min J, Xue H, Kaminsky LA, Cheskin LJ. Has the prevalence of overweight, obesity and central obesity levelled off in the United States? Trends, patterns, disparities, and future projections for the obesity epidemic. Int J Epidemiol. 2020; 1–14. doi:10.1093/ije/dyz273

2. Amugsi DA, Dimbuene ZT, Mberu B, Muthuri S, Ezeh AC. Prevalence and time trends in overweight and obesity among urban women: An analysis of demographic and health surveys data from 24 African countries, 1991 - 2014. BMJ Open. 2017;7: 1–12. doi:10.1136/bmjopen-2017-017344

3. Lin TC, Courtney TK, Lombardi DA, Verma SK. Association between Sedentary Work and BMI in a U.S. National Longitudinal Survey. Am J Prev Med. 2015;49: e117–e123. doi:10.1016/j.amepre.2015.07.024

4. World Health Organization. Global Status Report On Noncommunicable Diseases 2014. 2014.

5. Dean E, Söderlund A. What is the role of lifestyle behaviour change associated with non-communicable disease risk in managing musculoskeletal health conditions with special reference to chronic pain? BMC Musculoskelet Disord. 2015;16. doi:10.1186/s12891-015-0545-y

6. Ezzati M, Riboli E. Behavioral and dietary risk factors for non-communicable diseases. N Engl J Med. 2013;369: 954–964. doi:10.1056/NEJMra1203528

7. Barrense-Dias Y, Berchtold A, Akre C, Surís JC. The relation between internet use and overweight among adolescents: A longitudinal study in Switzerland. Int J Obes. 2016;40: 45–50. doi:10.1038/ijo.2015.146

8. Aghasi M, Matinfar A, Golzarand M, Salari-Moghaddam A, Ebrahimpour-Koujan S. Internet Use in Relation to Overweight and Obesity: A Systematic Review and MetaAnalysis of Cross-Sectional Studies. Adv Nutr. 2019;11: 349–356. doi:10.1093/advances/nmz073

9. Chaput JP, Després JP, Bouchard C, Tremblay A. The association between sleep duration and weight gain in adults: A 6-year prospective study from the Quebec Family Study. Sleep. 2008;31: 517–523. doi:10.1093/sleep/31.4.517

10. Lin PH, Lee YC, Chen KL, Hsieh PL, Yang SY, Lin YL. The relationship between sleep quality and internet addiction among female college students. Front Neurosci. 2019;13. doi:10.3389/fnins.2019.00599

11. Do YK, Shin E, Bautista MA, Foo K. The associations between self-reported sleep duration and adolescent health outcomes: What is the role of time spent on Internet use? Sleep Med. 2013;14: 195–200. doi:10.1016/j.sleep.2012.09.004

12. Eliacik K, Bolat N, Koçyiğit C, Kanik A, Selkie E, Yilmaz H, et al. Internet addiction, sleep and health-related life quality among obese individuals: a comparison study of the growing problems in adolescent health. Eat Weight Disord. 2016;21: 709–717. doi:10.1007/s40519-016-0327-z

13. Tsai MC, Strong C, Chen WT, Lee CT, Lin CY. Longitudinal impacts of pubertal timing and weight status on adolescent internet use: Analysis from a cohort study of taiwanese youths. PLoS One. 2018;13: 1–10. doi:10.1371/journal.pone.0197860

14. Tsitsika AK, Andrie EK, Psaltopoulou T, Tzavara CK, Sergentanis TN, Ntanasis-Stathopoulos I, et al. Association between problematic internet use, socio-demographic variables and obesity among European adolescents. Eur J Public Health. 2016;26: 617–622. doi:10.1093/eurpub/ckw028

15. Goel D, Subramanyam A, Kamath R. A study on the prevalence of internet addiction and its association with psychopathology in Indian adolescents. Indian J Psychiatry. 2013;55: 140–143. doi:10.4103/0019-5545.111451

16. Bener A, Bhugra D. Lifestyle and depressive risk factors associated with problematic internet use in adolescents in an Arabian Gulf culture. J Addict Med. 2013;7: 236–242. doi:10.1097/ADM.0b013e3182926b1f

17. Bradbury KM, Turel O, Morrison KM. Electronic device use and beverage related sugar and caffeine intake in US adolescents. PLoS One. 2019;14. doi:10.1371/journal.pone.0223912

18. Haque M, McKimm J, Sartelli M, Samad N, Haque SZ, Bakar MA. A narrative review of the effects of sugar-sweetened beverages on human health: A key global health issue. J Popul Ther Clin Pharmacol. 2020;27: e76–e103. doi:10.15586/jptcp.v27i1.666

19. Aryal KK, Mehata S, Neupane S, Vaidya A, Dhimal M, Dhakal P, et al. The burden and determinants of non communicable diseases risk factors in Nepal: Findings from a nationwide STEPS survey. PLoS One. 2015;10: 1–18. doi:10.1371/journal.pone.0134834

20. Rawal LB, Kanda K, Mahumud RA, Joshi D, Mehata S, Shrestha N, et al. Prevalence of underweight, overweight and obesity and their associated risk factors in Nepalese adults: Data from a nationwide survey, 2016. PLoS One. 2018;13: 1–14. doi:10.1371/journal.pone.0205912

21. Xu CX, Zhu HH, Fang L, Hu RY, Wang H, Liang M Bin, et al. Gender disparity in the associations of overweight/obesity with occupational activity, transport to/from work, leisure-time physical activity, and leisure-time spent sitting in working adults: A cross-sectional study. J Epidemiol. 2017;27: 401–407. doi:10.1016/j.je.2016.08.019

22. Poobalan A, Aucott L. Obesity Among Young Adults in Developing Countries: A Systematic Overview. Curr Obes Rep. 2016;5: 2–13. doi:10.1007/s13679-016-0187-x

23. Dixon LJ, Correa T, Straubhaar J, Covarrubias L, Graber D, Spence J, et al. Gendered Space: The digital divide between male and female users in internet public access sites. J Comput Commun. 2014;19: 991–1009. doi:10.1111/jcc4.12088

24. Su W, Han X, Jin C, Yan Y, Potenza MN. Are males more likely to be addicted to the internet than females? A meta-analysis involving 34 global jurisdictions. Comput Human Behav. 2019;99: 86–100. doi:10.1016/j.chb.2019.04.021

25. Amis N. Nepal Telecommunications Authority. 2019;174: 1–11. Available: https://nta.gov.np/wp-content/uploads/MIS-2076-Baisakh.pdf

26. Nepal Demographic and Health Survey 2016. Kutmundu; 2017.

27. Al Kibria GM. Prevalence and factors affecting underweight, overweight and obesity using Asian and World Health Organization cut-offs among adults in Nepal: Analysis of the Demographic and Health Survey 2016. Obes Res Clin Pract. 2019;13: 129–136. doi:10.1016/j.orcp.2019.01.006

28. Bishwajit G. Household wealth status and overweight and obesity among adult women in Bangladesh and Nepal. Obes Sci Pract. 2017;3: 185–192. doi:10.1002/osp4.103

29. Matusitz J, McCormick J. Sedentarism: The effects of internet use on human obesity in the United States. Soc Work Public Health. 2012;27: 250–269. doi:10.1080/19371918.2011.542998

30. Shrier I, Platt RW. Reducing bias through directed acyclic graphs. BMC Med Res Methodol. 2008;8: 1–15. doi:10.1186/1471-2288-8-70

31. Rana J, Ahmmad Z, Sen KK, Bista S, Islam RM. Socioeconomic Differentials in Hypertension based on JNC7 and ACC/AHA 2017 Guidelines Mediated by Body Mass Index: Evidence from Nepal Demographic and Health Survey. PLoS One. 2020;15. doi:10.1371/journal.pone.0218767

32. R Core Team. R: A language and environment for statistical computing. Vienna, Austria: R Foundation for Statistical Computing, Vienna, Austria.; 2020. Available: https://www.r-project.org/

33. Vandelanotte C, Sugiyama T, Gardiner P, Owen N. Associations of leisure-time internet and computer use with overweight and obesity, physical activity and sedentary behaviors: Cross-sectional study. J Med Internet Res. 2009;11: 1–8. doi:10.2196/jmir.1084

34. Afshin A, Forouzanfar MH, Reitsma MB, Sur P, Estep K, Lee A, et al. Health effects of overweight and obesity in 195 countries over 25 years. N Engl J Med. 2017;377: 13–27. doi:10.1056/NEJMoa1614362

35. Clement J. Internet usage worldwide - Statistics & Facts Statistica. 2015. Available: https://www.statista.com/topics/1145/internet-usage-worldwide/

36. Meral G. Is digital addiction a reason for obesity? Ann Med Res. 2018;25: 472. doi:10.5455/annalsmedres.2018.05.102

37. Chaput JP, Després JP, Bouchard C, Tremblay A. The association between sleep duration and weight gain in adults: A 6-year prospective study from the Quebec Family Study. Sleep. 2008;31: 517–523. doi:10.1093/sleep/31.4.517

38. Gaskin CJ, Orellana L. Factors associated with physical activity and sedentary behavior in older adults from six Lowand middle-income countries. Int J Environ Res Public Health. 2018;15. doi:10.3390/ijerph15050908

39. Kim JH, Lau CH, Cheuk KK, Kan P, Hui HLC, Griffiths SM. Brief report: Predictors of heavy Internet use and associations with health-promoting and health risk behaviors among Hong Kong university students. J Adolesc. 2010;33: 215–220. doi:10.1016/j.adolescence.2009.03.012

40. Matusitz J, McCormick J. Sedentarism: The effects of internet use on human obesity in the United States. Soc Work Public Health. 2012;27: 250–269. doi:10.1080/19371918.2011.542998

41. Rosiek A, Maciejewska NF, Leksowski K, Rosiek-Kryszewska A, Leksowski Ł. Effect of television on obesity and excess of weight and consequences of health. Int J Environ Res Public Health. 2015;12: 9408–9426. doi:10.3390/ijerph120809408

